# ImplemeNting SARS-CoV-2 Rapid antigen testing in the Emergency wArd of a Swiss univErsity hospital: the INCREASE study

**DOI:** 10.1101/2021.02.10.21250915

**Authors:** Giorgia Caruana, Antony Croxatto, Eleftheria Kampouri, Antonios Kritikos, Onya Opota, Marylin Foerster, René Brouillet, Laurence Senn, Reto Lienhard, Adrian Egli, Giuseppe Pantaleo, Pierre-Nicolas Carron, Gilbert Greub

**Author notes:** **Corresponding Author** Prof. Gilbert Greub, Institute Universitaire de Microbiologie, Département de médecine de laboratoire et pathologie (DMLP), Centre Hospitalier Universitaire Vaudois (CHUV); and Université de Lausanne, Rue du Bugnon 48, CH-1011, Lausanne, Switzerland. Personal.

## Abstract

**Background:** While facing a second wave in SARS-CoV-2 pandemic, in November 2020 the Swiss Federal Office of Public Health (FOPH) authorized the use of rapid antigen tests (RATs) in addition to the gold-standard reverse transcription-polymerase chain reaction (RT-PCR).

**Methods:** We implemented the use of RAT in the emergency ward of our university hospital for rapid patients’ triaging and compared performances of four different antigen tests. All results were compared to SARS-CoV-2 specific RT-PCR (reference standard).

**Results:** Triaging patients using RAT in association with RT-PCR allowed us to isolate promptly positive patients and to save resources, in a context of rapid RT-PCR reagents shortage. Among 532 patients with valid results, overall sensitivities were 48.3% for One Step Exdia and 41.2% for Standard Q^®^, Panbio−and BD Veritor. All four antigen tests exhibited specificity above 99%. Sensitivity increased up to 74.6%, 66.2%, 66.2% and 64.8% for One Step Exdia, Standard Q, Panbio, and BD Veritor respectively, when considering viral loads above 10^5^copies/ml, up to 100%, 97.8%, 96.6% and 95.6% for viral loads above 10^6^ copies/ml and 100% (for all tests) when considering viral loads above 10^7^ copies/ml. Sensitivity was significantly higher for patients presenting with symptoms onset within 4 days (74.3%, 69.2%, 69.2% and 64%, respectively) versus patients with evolution of symptoms for more than 4 days (36.8%, 21.1%, 21.1% and 23.7%, respectively). Sensitivities of all RAT assays were of only 33% among hospitalized patients without COVID-19 symptoms.

**Conclusion:** RAT might represent a useful epidemiological resource in selected clinical settings as a complementary tool to the molecular tests for rapid patients triaging, but the lower sensitivity compared to RT-PCR, especially in late presenters and subjects without COVID-19 symptoms, must be taken into account in order to correctly use RAT for triaging.

## Introduction

Since the beginning of the outbreak in Wuhan (China) at the end of 2019^1^ and through the rapid transformation into a pandemic^2^, SARS-CoV-2 has inflicted tremendous changes to the public health management. Countries all over the world had to adapt their priorities, constantly increasing the number of tests performed and shortening the time to results, in order to follow the chains of transmissions and to promptly cohort infected patients.

During November 2020, the City of Lausanne (Vaud canton, Switzerland), was one of the areas with the highest SARS-CoV-2 positive rates worldwide, with about 1800 positive new cases per/100.000 inhabitants in 14 days^3^. During this time and for several weeks, Lausanne University hospital (CHUV), a tertiary-care center of 1500 beds, experienced a massive influx of patients, rapidly exceeding its overall capacity, with up to 280 patients hospitalized for COVID-19, including 56 requiring Intensive-Care-Unit (ICU) management.

Among patients consulting in the emergency department, approximately 50% (around 20 patients per day) presented symptoms compatible with acute COVID-19, thus necessitating preliminary and rapid triaging in order to facilitate their timely isolation and orientation in dedicated units and to ensure the safety of other patients. Our emergency service acts as a university reference center, but also as a primary care hospital for the inhabitants of the region of Lausanne.

From November 2, the Swiss Federal Office of Public Health (FOPH) authorized the use of rapid antigen tests (RATs), in addition to the gold-standard real-time reverse transcription-polymerase chain reaction (RT-PCR), mainly for outpatients in testing centers, pharmacies and medical private practices, aiming to broaden screening access^4^. This decision was based on an official recommendations from the members of the Coordination Commission of Clinical Microbiology (CCCM) of the Swiss Society of Microbiology (SSM)^5^. According to these recommendations, the use of RAT is indicated for i) patients with acute respiratory symptoms during less than 4 days, not requiring hospitalization, or ii) early triage of patients to be hospitalized, in a context of epidemiological emergencies (such as an outbreak), with an high number of patients admitted per day and a pre-test probability above 20%^5^.

Among the multiple antigen tests commercially available^6^, the chosen reference standard antigen tests were the Standard Q^®^ COVID-19 Rapid Antigen Test (SD Biosensor-Republic of Korea /Roche - Switzerland) and the Panbio−COVID-19 Ag Rapid Test (Abbott - USA). These were validated in two clinical studies carried out in Switzerland, one at the University Hospital of Geneva (HUG, Geneva, Switzerland) and the other one between the CHUV (at our diagnostic laboratory) and the University Center of General Medicine and Public Health (Unisanté, Lausanne, Switzerland)^7,8^. Both antigen tests showed high performances (respectively 85% and 87.4% of sensitivity and above 99% of specificity) among patients with recent infection (less than 7 days of symptoms) consulting outpatient testing centers in Geneva and Lausanne.

On the 7^th^ of November 2020, we implemented the diagnostic flow of SARS-CoV-2 infection at our hospital with a new RAT laboratory enforced in the heart of the emergency department, in order to reduce delivery time, thus achieving early placement of positive patients in COVID dedicated units and, as secondary benefit, a reduction of rapid RT-PCR assays.

During this implementation, we also compared performances of the reference standard tests from Abbott and Roche with two other antigen tests. These additional tests were chosen for their advantage to be fully automated, owing to their automatic reading system with a possible direct connection between the reads and the laboratory interface system (LIS), thus allowing a potential reduction in the turn-around time (TAT), human error rate, as well as the need of technical expert personnel. The performances of all four tests were stratified according to different cycle-thresholds (Ct)/ viral loads (VL) and the delay since symptom onset.

Furthermore, because of the inherent characteristics of the population admitted to the emergency department of a University hospital, with usually more patients severely ill/or with longer duration of illness, in comparison to the outpatient population consulting in testing centers and clinics, we formulated the hypothesis that the presence of nasopharyngeal IgA might influence the sensitivity of RATs, potentially masking the antigen-targets of RATs. IgA is a key component in humoral immune response, thus abundant in external secretions^9^. Several data in the literature already demonstrated the early production of IgA in response to SARS-CoV-2 infection, starting from the first week after the infection^10,11^. For this reason, we measured nasopharyngeal IgA among patients tested with antigen kits and we compared the medians of Ct values between IgA positive and IgA negative patients.

## Methods

### Laboratory set up, type of antigen tests and testing procedures

A specific RAT laboratory was built inside the emergency department, with two laboratory technicians dedicated working there 8 hours per day (from 9:00 to 18:30, given the highest activity during these hours), 7 days out of 7, receiving nasopharyngeal samples taken from every patient consulting the ER from November 7.

The nasopharyngeal swabs were delivered from the patient to the RAT laboratory, immediately after the sampling procedure and their labelling with the patient’s demographic details, hospital code and a red (COVID-19 symptomatic) or green (COVID-19 asymptomatic) label to quickly cohort patients according to our diagnostic algorithm (Fig. 1).

**Figure 1.**
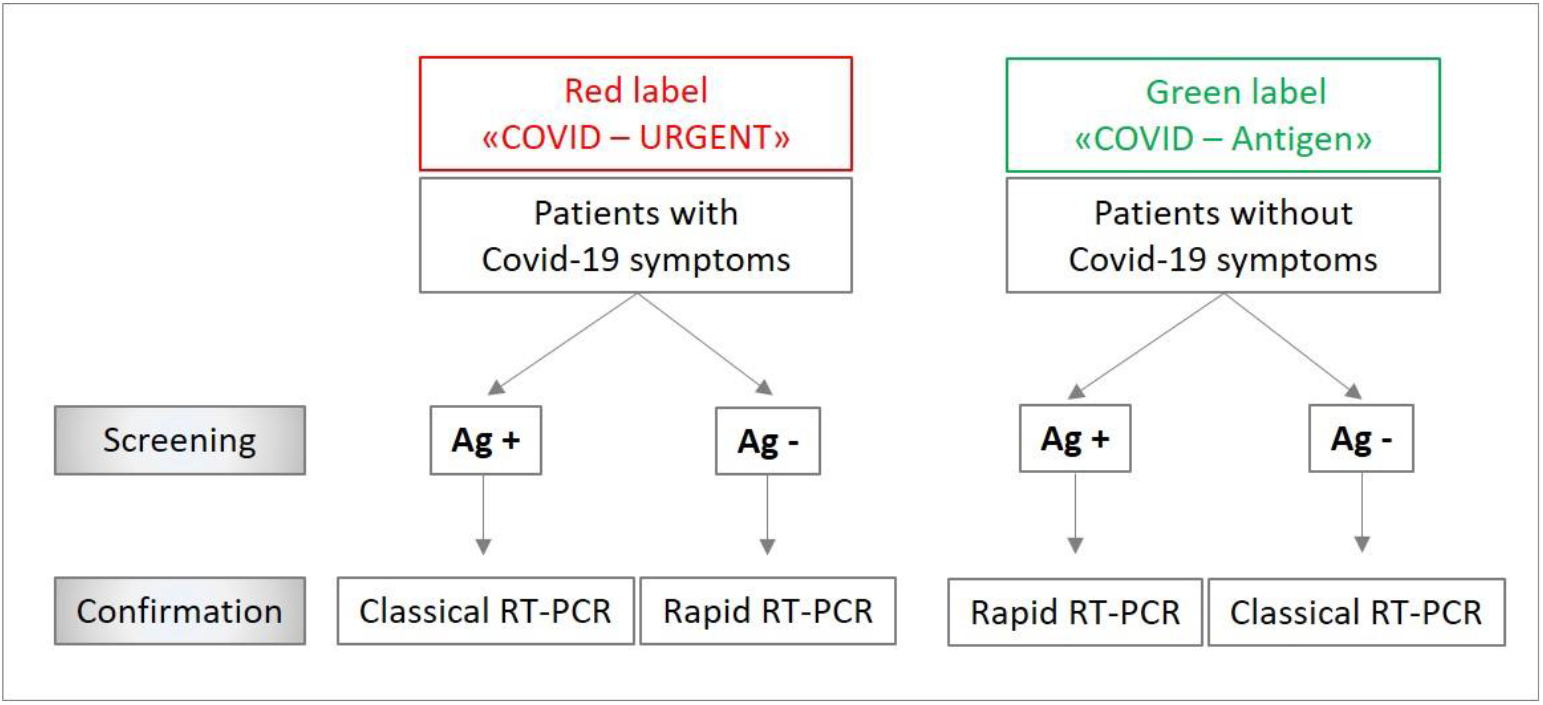
Diagnostic algorithm for managing tests flow according COVID-19 symptoms. Ag +: positive rapid antigen test. Ag-: negative rapid antigen test. RT-PCR: real-time reverse transcription-polymerase chain reaction.

Nasopharyngeal swabs were analyzed using four different kits: Standard Q^®^ COVID-19 Rapid Antigen Test (SD Biosensor - Republic of Korea /Roche - Switzerland), Panbio−COVID-19 Ag Rapid Test (Abbott - USA), One Step Immunoassay for Exdia COVID-19 Ag (Precision Biosensor Inc. - Republic of Korea) and The BD Veritor™ System for Rapid Detection of SARS-CoV-2 (Becton Dickinson, USA). Standard Q^®^ COVID-19 Rapid Antigen Test was considered the reference standard for RATs and its results were used for patients care. With the purpose of performing RT-PCR confirmation on every sample avoiding patients’ discomfort due to double sampling, the evaluation was done using a wet swab procedure, by suspending the nasopharyngeal swabs in 2.5 to 3 ml of viral transport media (VTM) solution. Then, 300 µl (for Panbio−, BD Veritor™ and One Step Immunoassay) or 350 µl (for Standard Q^®^) of the sample were mixed with the buffer solution and then tested, according to the manufacturer instructions (Fig. 2). The incubation and the reading of the BD Veritor™ and One Step Immunoassay tests was performed automatically with the provided reader after 20 minutes, while the reading of the Standard Q^®^ and Panbio−tests was carried out visually by the laboratory technician after 15 to 30 minutes of incubation.

**Figure 2.**
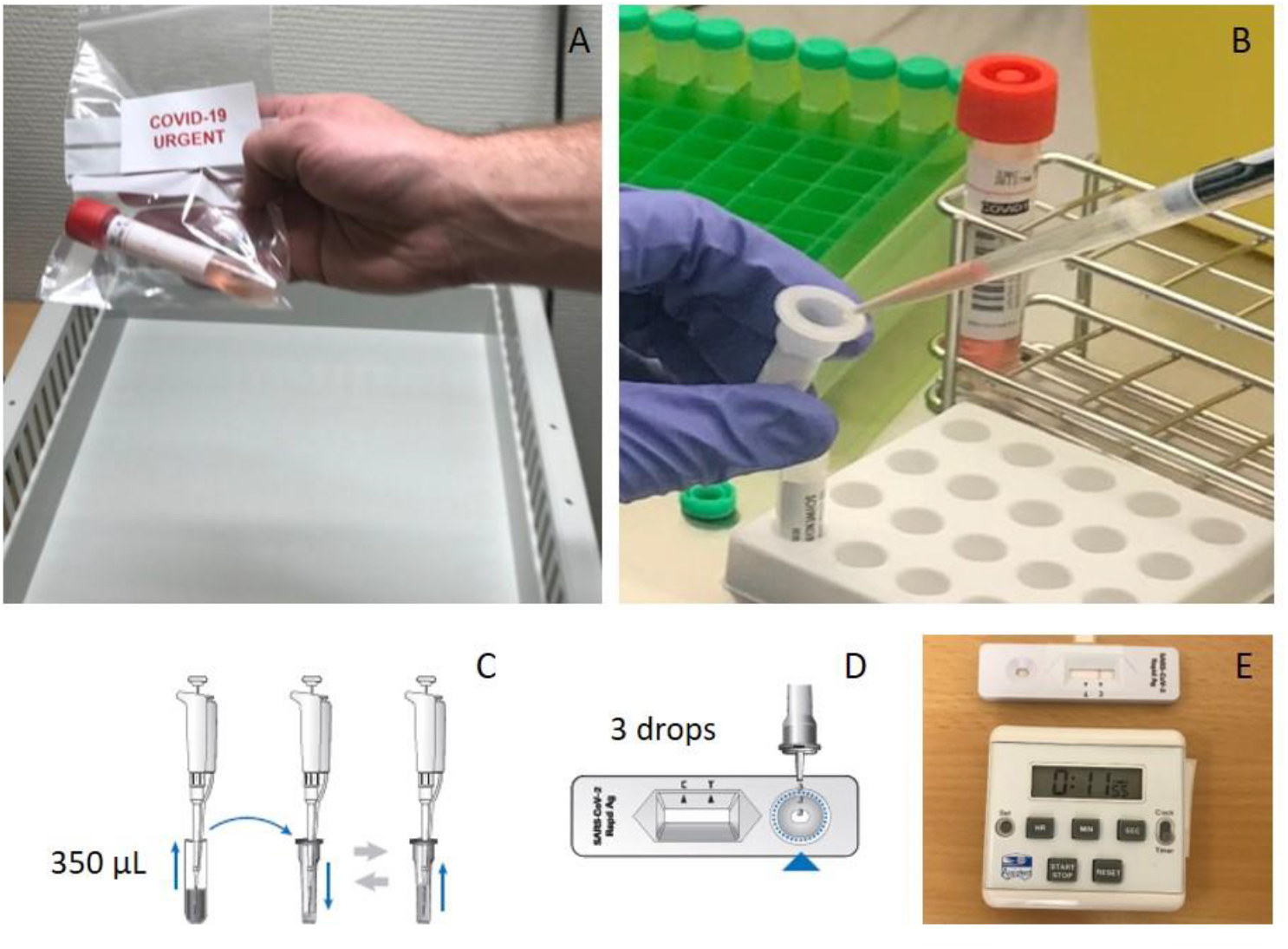
Antigen testing procedure with reference test (Roche). A. Fresh nasopharyngeal sample from a COVID-19 symptomatic patient received at the RAT laboratory. B. 350 microliters of sample are collected from the viral transport medium and (C) mixed with the extraction buffer, according to manufacturer instructions. D. Three drops of extracted sample are applied into the testing device. E. Results are readable in 15 to 30 minutes.

### Routine confirmation by RT-PCR

Given the uncertainty regarding sensitivity and specificity of RAT, all antigen results were confirmed on one of the following molecular platforms: i) VIASURE SARS-CoV-2 (N1 + N2) Real Time PCR Detection Kit for BD MAX™ (Becton Dickinson, USA) or GeneXpert SARS-CoV-2 test (Cepheid, https://www.cepheid.com)^12^ as rapid systems, ii) test cobas 6800® SARS-CoV-2 (Roche, Basel, Switzerland)^13^ or our automated high-throughput molecular diagnostic (MDx) platform as classic systems. Our MDx platform uses Magnapure RNA-extraction coupled to applied biosystems 7900 amplification device (Quant Studio 7) and three Hamilton robots, with primers targeting the E- and RdRp-encoding genes^14,15^. The RT-PCR system (rapid RT-PCR versus classical RT-PCR) was chosen according to a diagnostic algorithm (Fig. 1) in order to facilitate patient flow and optimize the use of rapid PCR systems, without compromising patient’s clinical management.

### Study design of RAT comparison

The performance comparison study of antigen tests was carried out on a prospective collection of upper respiratory specimens from all patients presenting at the emergency unit of CHUV. All patients admitted to the hospital wards, intermediate care units or ICU were systematically screened for SARS-CoV-2. All patients consecutively admitted to the hospital from the emergency department, with or without COVID-19 signs or symptoms, were thus included in the study. The same samples were tested and confirmed as described above. Patients’ enrollment for RAT comparison was stopped once we reached a collection of 100 PCR positive samples and at least 200 PCR negative samples. The minimum targeted sample size was calculated according to the SSM recommendations for RAT validation^5^.

Clinical history (duration and type of symptoms), demographic details and nasopharyngeal specimens were collected for each patient as part of standard of care. Microbiological data and time to results were extracted from our LIS.

### Statistical analysis

In order to quantify the viral load (VL) based on the number of Ct obtained with different molecular platforms, we used the following equation, derived from RNA quantification: VL=(10^((Ct −40.856)/ −3.697))*100. Details on methods used to derive this equation were described elsewhere^16^. Briefly, a plasmid with the target sequence of the PCR (RD-Biotech, Besançon, France) or purified viral RNA were used to convert Ct obtained on our MDx platform to VL. We demonstrated a proportional bias in the inter-variability of Ct measures between VIASURE SARS-CoV-2 (N1 + N2) Real Time PCR Detection Kit for BD MAX™ and GeneXpert SARS-CoV-2 test compared to cobas 6800® SARS-CoV-2 through Bland-Altman analyses (Supplementary material, Tab. 1). In order to adjust for this bias, Passing-Bablock regression equations were used to re-adapt BD-MAX (Ct calculated= 0.8829*Ct + 4.882) and GeneXpert Ct values (Ct calculated= 4.38 + 0.9*Ct if Ct > 20, or Ct calculated= 3.85 + 0.92*Ct if Ct< 20) Ct (Supplementary material, Fig. 1).

**Table 1.** Estimated disease prevalence, sensitivity and specificity according to different diagnostic approaches, in subjects with or without symptoms of COVID admitted at the emergency ward of Lausanne University Hospital. Please, note that in this setting, the RAT only detected about one third of asymptomatic hospitalized patients, who resulted positive by RT-PCR.

|  | **Without COVID-19**<br>**symptoms**<br>**(N=239)** | **With COVID-19**<br>**symptoms**<br>**(N=293)** | ***p*-value** |
| --- | --- | --- | --- |
| **Gender** |  |  |  |
| Female | 105 (43.9%) | 131 (44.7%) | 0.927 |
| Male | 134 (56.1%) | 162 (55.3%) |  |
| **Age** |  |  |  |
| Median [IQR] years | 67.0 [48.5, 81.0] | 75.0 [61.0, 85.0] | <0.001 |
| **SARS-CoV-2 RT-PCR result** |  |  |  |
| Negative | 215 (90.0%) | 203 (69.3%) | <0.001 |
| Positive | 24 (10.0%) | 90 (30.7%) |  |
| **SARS-CoV-2 RAT\* result** |  |  |  |
| Negative | 231 (96.7%) | 253 (86.3%) | <0.001 |
| Positive | 8 (3.3%) | 40 (13.7%) |  |
| **Cycle threshold** |  |  |  |
| Median [IQR] | 29.8 [22.6, 35.1] | 27.0 [20.5, 32.5] | 0.274 |
| Missing | 215 (90.0%) | 203 (69.3%) |  |

\*RAT results represented here were obtained with Standard Q (Roche) test, which was used for patients care. RAT: rapid antigen detection. RT-PCR: real-time reverse transcription-polymerase chain reaction. IQR : Interquartile range.

We evaluated detection rate, positive and negative predicted value (PPV, NPV), sensitivity, and specificity of each test with 95% confidence intervals (CIs) using exact binomial test and a one-sided test; in each case, overall accuracy and Kappa statistics were calculated. Results from RT-PCR were used as reference for sensitivity and specificity calculations. Analyses were stratified by viral load categories and by time delay of symptoms presentation. Both Ct and different time delay categories (before day 4, between day 4 and 7, after day 7) were chosen based on recent data in the literature^8,17^.

Finally, Wilcoxon rank sum test with continuity correction and Chi-squared test were performed to compare continuous and categorical variables respectively, when appropriate.

Data were analyzed using “mcr”, “blandr”, “caret” and “table1” packages on “R statistical software” (version 3.6.1, 2019, Vienna, Austria).

### Ethical declaration

This article was prepared according to STANDARD guidelines for diagnostic accuracy studies reporting. The data on the fiability of the different antigen assays were obtained during a quality enhancement project at our institution (CHUV, Lausanne). According to national law (Swiss Federal Act on Human Research), the performance and publishing the results of such a project can be done without asking the permission of the competent research ethics committee

## Results

### Emergengy department patients flow

From November 7 to December 16, all patients presenting at the emergency department, with or without suspected of SARS-CoV-2 infection, were tested with at least one antigen test at the laboratory built inside the emergency ward at Lausanne University Hospital (CHUV). Standard Q antigen test from Roche was used as reference, thus only its result was available to patients and clinicians. A total of 572 patients were screened; among 532 patients with available results, 293 (55.1%) had symptoms consistent with COVID-19 and 239 (44.9%) were admitted for other reasons than COVID-19 suspicion (Fig. 3). Patients with symptoms of COVID −19 were significantly older (*p*<0.001) than asymptomatic ones, with a median age of 75 [IQR: 61-85] and 67 [IQR: 48.5-81] years, respectively (Tab. 1). No significant differences were found between the proportions of symptomatic men (162, 55.3%) and women (131, 44.7%, Tab. 1). As expected, there was a significant difference (*p*<0.001) between the proportion of positive RAT results between symptomatic (40/293, 13.7%) and asymptomatic (8/239, 3.3%) patients, which was corroborated by the RT-PCR results (90/293, 30.7% versus 24/239, 10%), while no significant differences were found between the median Ct of symptomatic versus non-symptomatic patients (Tab. 1).

**Figure 3.**
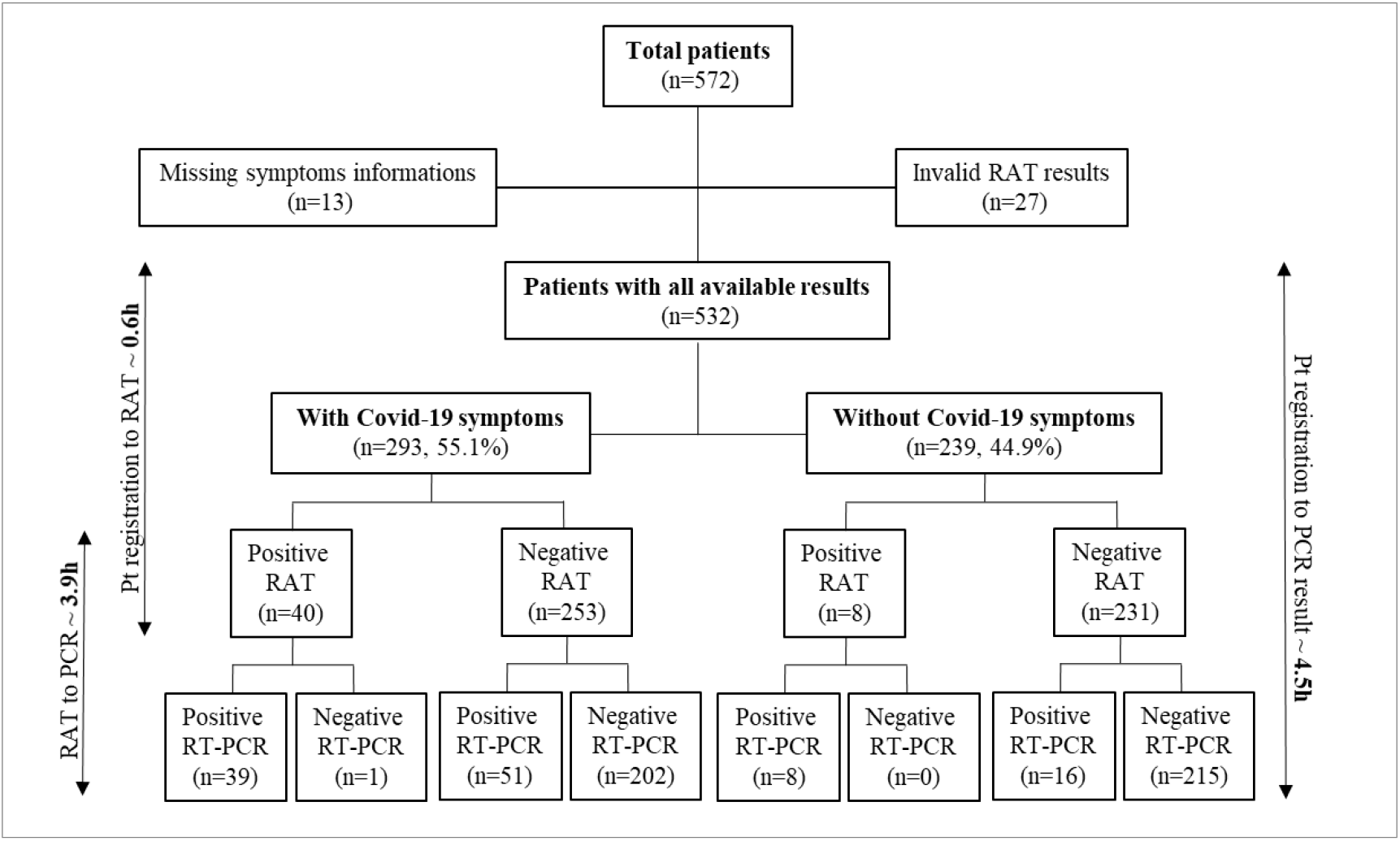
Number of patients included and time-to-results from patients’ registration to RAT or RT-PCR result. RAT: Rapid Antigen Test. RT-PCR: reverse transcription-polymerase chain reaction. Pt: patients. H : hours. Please, note that the time-to-result analysis was performed on available results from 375 patients.

According to our diagnostic algorithm (Fig. 1), patients with symptoms of Covid-19 who tested positive with the antigen test and patients without symptoms of Covid-19 who tested negative with antigen test were confirmed with one of our classical RT-PCR platforms. The remaining patients, showing discordant results compared to their clinical presentation, were confirmed using one of our rapid molecular platforms. All symptomatic patients with negative antigen results stayed at the emergency department in an isolation room or were admitted in a dedicated isolation unit until the result of RT-PCR. Overall, this strategy allowed us to save 271 rapid RT-PCR tests.

Among all patients submitted to RAT, we documented one false positive result and 67 false negative results (51 among COVID-19 symptomatic patients and 16 among COVID-19 asymptomatic patients, as shown in Fig. 3). The false positive result belonged to a patient admitted for worsening of dyspnea, cough and weakness in the last three days. Because of the concordance between clinical presentation (suspicion of SARS-CoV-2 infection) and the result of antigen test, the patient was cohorted in a room with other COVID-19 positive patients. Thanks to our diagnostic flow with RT-PCR confirmation, we were able to identify the discordance and move the patient to another room as soon as the molecular results were available. In order to exclude possible cross-reactions of the antigen test with other respiratory pathogens, a multiplex PCR panel (testing *Chlamydia pneumoniae, Mycoplasma pneumoniae*, Adenovirus, Parainfluenza virus 1 to 4, Human metapneumovirus, Pan-entero/Rhinovirus, Coronavirus E229, OC43, HKU1, NL63) was performed, which all resulted negative. The patient remained free of COVID-19 symptoms after discharge and subsequent testing performed over 14 days later remained negative.

### Diagnostic test performance of RATs

A total of 572 patients were consecutively tested with the four RAT systems: One Step Exdia, Standard Q, Panbio, and BD Veritor. All antigen test results were initially confirmed as described above with one of our four molecular platforms. Among 532 valid results, we obtained an overall sensitivity 48.3%, a PPV of 96.5% and a NPV of 87.6% for One Step Exdia, an overall sensitivity of 41.2%, a PPV of 97.9% and a NPV of 86.2% for Standard Q, values of 41.2%, 95.9% and 86.1%, respectively, for BD Veritor and of 41.2%, 97.9% and 86.2% respectively for Panbio test (Tab. 2). Specificity was greater than 99% for all the antigen tests (Tab. 2). Sensitivity results increased up to 74.6%, 66.2%, 66.2% and 64.8% for One Step Exdia, Standard Q, Panbio, and BD Veritor respectively, among 71 patients with VLs greater than 10^5^, up to 100%, 97.8%, 96.6% and 95.6% among 46 patients with VLs greater than 10^6^ and up to 100% (for all tests) when considering 38 patients with VLs greater than 10^7^ (Fig. 4 and supplementary material, Tab. 2).

**Table 2.** Estimated disease prevalence, sensitivity and specificity according to different tests.

| Type of RAT | Prevalence with RT-PCR | Prevalence with RAT | Overall sensitivity of RAT | Overall specificity of RAT | RAT accuracy [95% CI] |
| --- | --- | --- | --- | --- | --- |
| One Step | 114/532 (21.4%) | 57/532 (10.7%) | 48.3% | 99.5% | 88.5% [85.5-91.1] |
| Standard Q | 114/532 (21.4%) | 48/532 (9%) | 41.2% | 99.7% | 87.2% [84.1-89.9] |
| Panbio | 114/532 (21.4%) | 49/532 (9.2%) | 41.2% | 99.5% | 87.0% [83.9-89.8] |
| BD Veritor | 114/532 (21.4%) | 48/532 (9%) | 41.2% | 99.7% | 87.2% [84.1-89.9] |

RAT: rapid antigen detection. RT-PCR: real-time reverse transcription-polymerase chain reaction.

**Figure 4.**
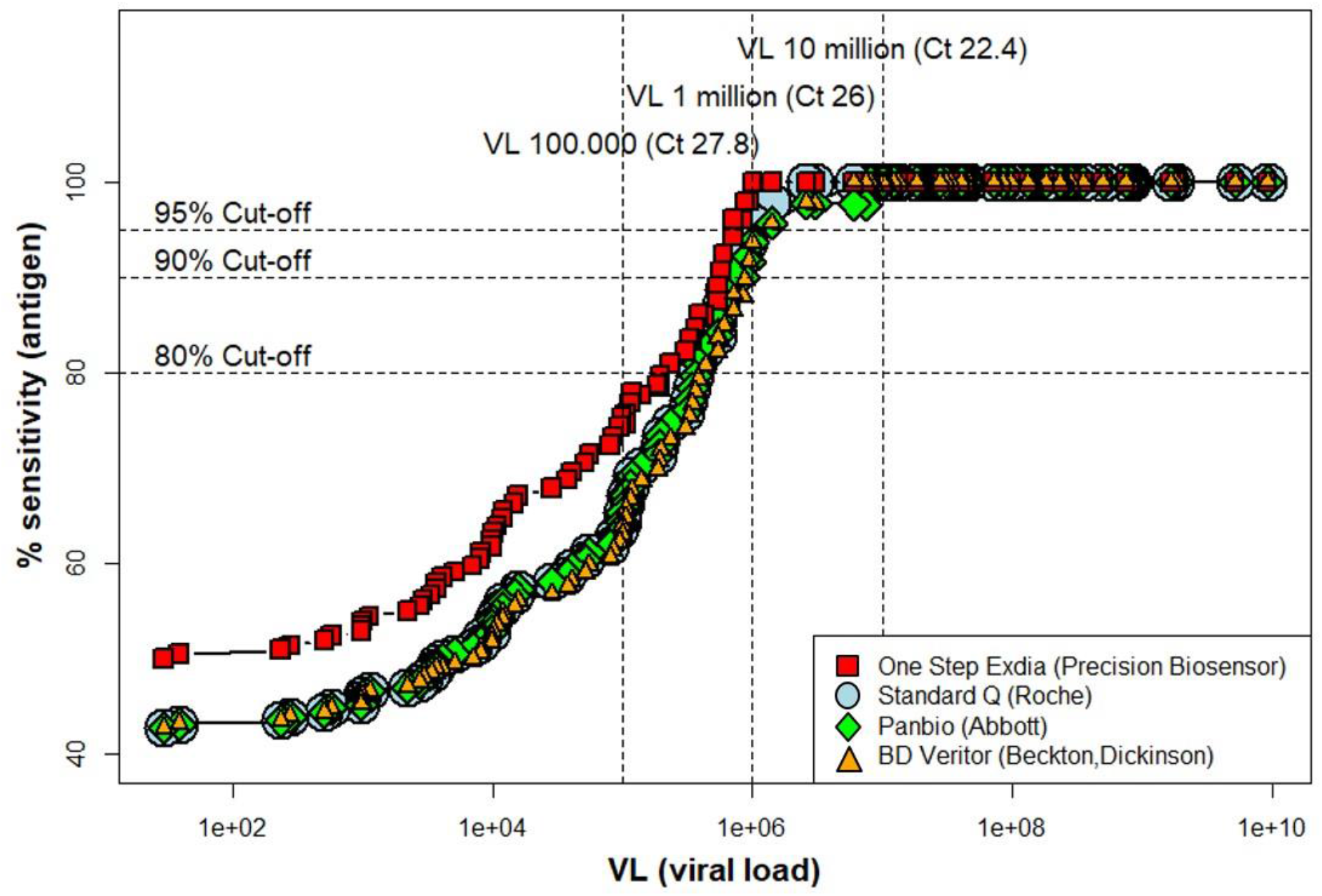
Comparison between four rapid antigen tests, showing the sensitivity according to the viral load. Please, note that the One Step Exdia test exhibited the best performances, with 75.9% sensitivity for VL>10^5^ copies/ml and 100% sensitivity for VL>10^6^ and 10^7^ copies/ml.

A clear difference was observed when comparing patients with and without symptoms of COVID-19, with overall sensitivities of 33% for all antigen tests for patients without (Tab. 3).

**Table 3.** Overall sensitivity and specificity rates according to different tests, symptoms onset delay and compared between COVID-19 symptomatic and asymptomatic patients.

|  | Se./Sp. rates<br>(Without COVID-19 symptoms) | Se./Sp. rates<br>(With COVID-19 symptoms) |  |  |  |
| --- | --- | --- | --- | --- | --- |
| RAT | Overall<br>(n=239) | Overall<br>(n=293)* | Symptoms onset delay ≤4 days<br>(n=138) | Symptoms onset delay 4≤7 days<br>(n=46) | Symptoms onset delay >7 days<br>(n=44) |
| One Step |  |  |  |  |  |
| - Se. | 33% | 52.2% | 74.3% | 43.7% | 31.8% |
| - Sp. | 100% | 99% | 100% | 96.7% | 95.4% |
| Standard Q |  |  |  |  |  |
| - Se. | 33% | 43.3% | 69.2% | 25% | 18.2% |
| - Sp. | 100% | 99.5% | 98.9% | 100% | 100% |
| Panbio |  |  |  |  |  |
| - Se. | 33% | 43.3% | 69.2% | 25% | 18.2% |
| - Sp. | 99.5% | 99.5% | 98.9% | 100% | 100% |
| BD Veritor |  |  |  |  |  |
| - Se. | 33% | 43.3% | 64% | 37.5% | 13.6% |
| - Sp. | 100% | 99.5% | 100% | 100% | 95.4% |

\*For 65/293 subjects labelled as COVID-19 symptomatic, either data were missing on symptoms onset delay, or patients did not show typical COVID-19 symptoms (differential diagnosis was not in favor of COVID-19).
RAT: rapid antigen test. Se.: sensitivity. Sp.: sensitivity.

Variations in overall sensitivity were observed also when accounting for patients’ duration of symptoms (Fig. 5 – Panels A-B and Tab. 3). Considering patients with a symptoms delay no longer than 4 days, we observed significantly higher sensitivity values of 74.3%, 69.2%, 69.2% and 64% for One Step Exdia, Standard Q, Panbio, and BD Veritor respectively, as compared to subjects with COVID-19 symptoms longer than 4 days (36.8%, 21.1%, 21.1% and 23.7%, respectively). Specifically, we obtained sensitivity rates of 43.7%, 25%, 25%, 37.7% respectively when considering patients with symptom duration between 4 and 7 days, further declining to 31.8%, 18.2%, 18.2% and 13.6% respectively for patients with symptoms longer than 7 days, in both cases with some sub-optimal specificity values (Tab. 3). To investigate the correlation of symptoms duration and variations in viral load, we also assessed these variables among patients admitted at CHUV from January to June 2020, during the first wave of SARS-CoV-2 infection. Data gathered from 444 patients showed a progressive reduction in viral load over time, with median Ct starting around 22 for the first 4 days since the symptoms onset, increasing to a median of 25 for symptoms delay between day 5 and 7 and finally up to a median Ct of 32 when considering symptoms dated longer than 7 days (Fig.2, supplementary material). A statistically significant difference was observed between median Ct before and after 4 days of symptoms delay (*p*<0.01).

**Figure 5.**
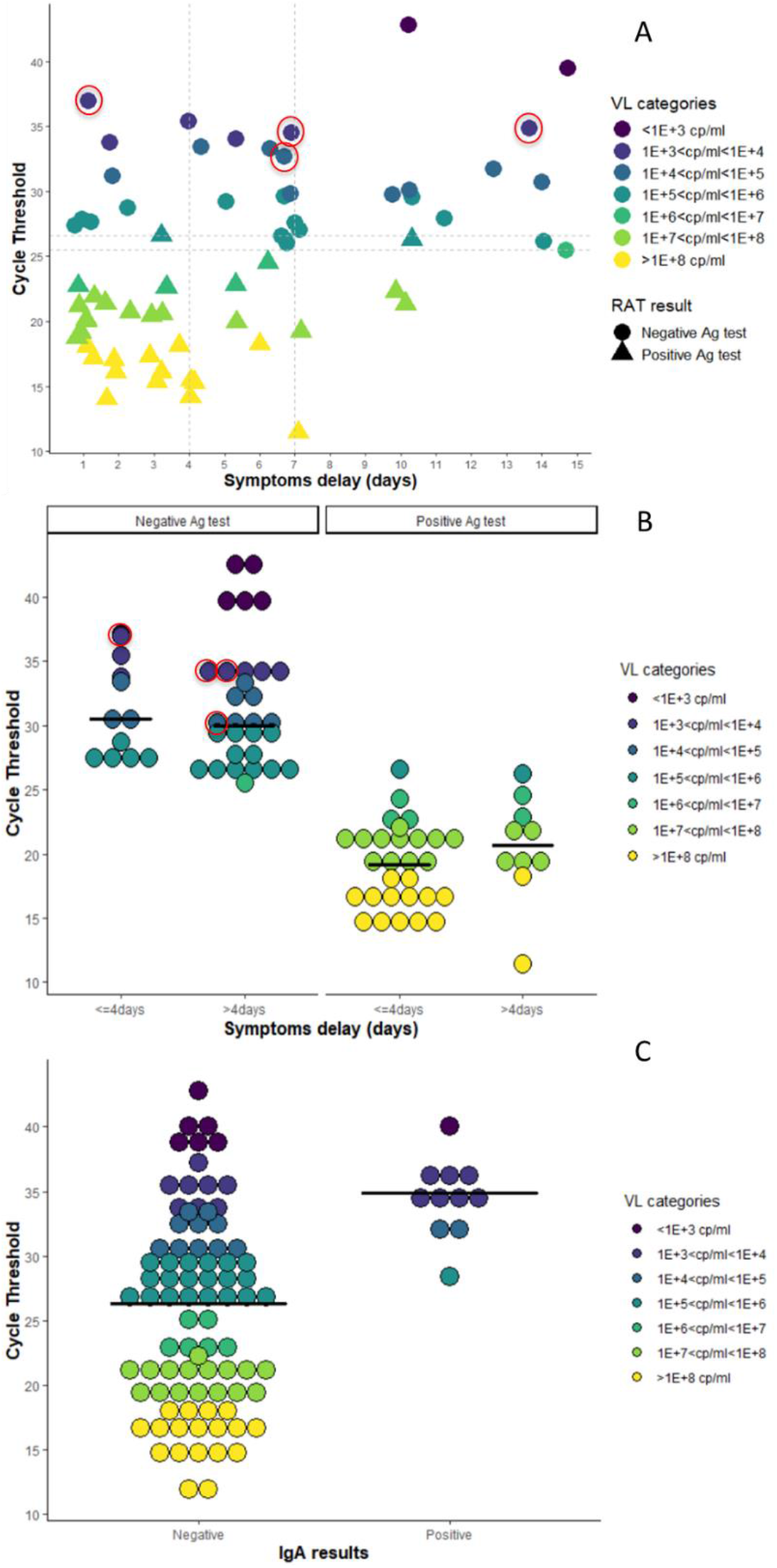
Cycle thresholds and viral loads of patients according to duration of symptoms and presence or absence of nasopharyngeal IgA. (A-B) Red circles represents samples tested positive for nasopharyngeal IgA. (B) Median Ct among positive and negative antigen tests according to symptoms duration. The test represented here is the Standard Q (Roche). (C) Viral load expressed in Ct according to the nasopharyngeal IgA result: please note that none of the 10 patients with positive IgA result had a positive RAT.

Notably, when compared to RT-PCR, RAT underestimated the prevalence of the disease to 9-10.7%, compared to the true prevalence of 21.4% as determined by RT-PCR (Tab. 2).

### Time-to-results evaluation

We finally performed an assessment of the time-to-results between i) the patient’s registration on our informatics system, ii) the result of the RAT and iii) the RT-PCR confirmation (final diagnosis). Among 375 patients for whom time to results was available, a mean of 0.6 hours (SD ± 1.8) since the time of patients’ registration was needed to obtain the result of antigen test, as compared to a mean of 4.5 hours (SD ± 6.4) for the result of RT-PCR; a mean delay of 3.9 hours (SD ± 6.8) was observed between the result of antigen test and the one of RT-PCR (Fig. 3).

### Nasopharyngeal IgA

We tested 95 patients for nasopharyngeal mucosal IgA: among them, 11/95 (11.6%) showed positive IgA levels (6 COVID-19 symptomatic and 5 COVID-19 non-symptomatic patients) and all of them were tested negative for RAT. Conversely, all 41 patients tested positive with RAT resulted negative for nasopharyngeal IgA, suggesting a possible implication of IgA in the competition with the antibodies used in the antigen assay. Remarkably, patients with nasopharyngeal IgA exhibited significantly higher median Cts (34.9, IQR 33.2-35.6) compared to those without IgA (26.3, IQR 19.2-30.7, *p*<0.001, Panel C, Fig. 5), thus further contributing to false negative RATs.

## Discussion

In this study, we describe how to implement and use RAT in an emergency room. Overall, as a consequence of the change in our workflow, by triaging patients through RAT results and by installing a RAT laboratory directly inside the emergency department, not only we were able to identify and isolate COVID-19 symptomatic and RAT positive patients within an average of 40 minutes from their registration, but we also succeeded in saving around 50% of the reagents for rapid molecular systems during this period. This management proved to be a precious resource, especially in a context of reagents shortage such as the one of a rapidly evolving pandemic. Short time to results might also have played a pivotal role in early placement of SARS-CoV-2 positive patients into COVID units, thus reducing risks of cross-transmission in emergency department. We also carried out a performance assessment and comparison between four antigen tests. Our results showed a significantly lower overall detection rate of SARS-CoV-2 infections compared to the previous validation we performed in Lausanne, and compared to other studies^8,18,19^. Because of the best performances of One Step Immunoassay from Exdia and the more convenient automatized reading method, once we terminated the comparison and validated the test, we decided to adopt this test as reference at our hospital.

Considering an acceptable sensitivity rate above 80%, none of the antigen tests in our study reached that threshold among patients with viral loads below 10^6^ copies/ml; sensitivity rates became good and excellent with viral loads above 10^6^ copies/ml (Tab. 4). Data from a recent epidemiological study on SARS-CoV-2 epidemics onset in Switzerland (Ladoy A. *et al*, manuscript in preparation) demonstrated the possible onset of clusters of infections originated from patients with VL below 10^6^ copies/ml, thus indirectly highlighting the risk associated with a massive (only-) RAT screening, which would likely lead to new and larger cases clusters in the population.

**Table 4.** Relationship between viral load, hypothetical contagiousness and correspondence with the sensitivity of diagnostic tests (based on our evaluation).

| Cycle threshold<br>(Ct, number) | Viral Load<br>(VL, copies/ml) | Hypothetical<br>contagiousness<sup>§</sup> | Sensitivity<sup>°</sup> of<br>RAT | Sensitivity<sup>°</sup><br>of RT-PCR |
| --- | --- | --- | --- | --- |
| >37.2 | <10<sup>3</sup> | Negligible | 0% | Acceptable |
| 37.2 – 33.5 | 10<sup>3</sup>-10<sup>4</sup> | Very low | 0% | Good |
| 33.5 – 29.8 | 10<sup>4</sup>-10<sup>5</sup> | Significant* | Very low* | Excellent |
| 29.8 – 26.1 | 10<sup>5</sup>-10<sup>6</sup> | High* | Low* | Excellent |
| 26.1 – 22.4 | 10<sup>6</sup>-10<sup>7</sup> | High | Acceptable | Excellent |
| 22.4 – 18.7 | 10<sup>7</sup>-10<sup>8</sup> | High | Excellent | Excellent |

RAT: rapid antigen detection. RT-PCR: real-time reverse transcription-polymerase chain reaction.
<sup>°</sup>Very low 30 to 60%; low 60 to 80%; acceptable > 80%; good > 95%; excellent > 99%
\*Critical area for which the antigen test is insufficient to detect important cases.
<sup>§</sup>Contagiousness is estimated to be negligible when $R_0 < 0.01$ ; very low when $R_0 > 0.01$ & $< 0.05$ ; significant when $R_0 > 0.05$ & $< 0.2$ ; high when $R_0 > 0.2$ over a 24 hour period in the same room than another patient.

We identified important differences between tests sensitivities according to the time of symptoms onset, with sensitivities dropping significantly starting from a “symptoms delay” greater than 4 days (Tab. 3). We hypothesized that the lower sensitivity was mainly due to lower viral load after 4 days of disease, and the results from studying the variation in viral loads among patients admitted to CHUV during the first wave of infections corroborated this hypothesis, showing a progressive reduction in viral load over time. The very low sensitivity (13.6%-31.8%, Tab. 3) of antigen tests in subjects with symptoms for more than 7 days is likely due to the low viral load generally observed after a week of disease in immunocompetent subjects (Supplementary fig. 2). To date, according to the official recommendations from the FOPH, RAT testing is indicated only within the first 4 days of symptoms^4^. Our results corroborated the importance of following this “less than 4 days symptoms delay” guideline for RAT screening, in order not to further decrease the test performances.

Notwithstanding the enthusiasm for rapid, cheaper and easy diagnostic solution such as antigen tests, the comparison between the sensitivity of RAT to the one of RT-PCR (Tab. 4) still highlights important differences in detection rates. It becomes clear that if we only used RAT, not only the prevalence of the disease would have been underestimated, but also a significant amount of patients with a high VL (from 10^4^ to 10^6^ copies/ml and thus potentially contagious) would not have been identified and therefore not put on isolation measures in a hospital with majority of 2 to 5-bed rooms. This is even more relevant among hospitalized patients with important comorbidities (hence more likely to develop complications), where appropriate cohorting would mean that fewer, potentially susceptible patients would be exposed for a prolonged duration to SARS-CoV-2.

Unfortunately, molecular systems are not available in all centers and, in consideration of the rapidly evolving epidemiological situation, contact tracing and early testing have become pivots for infection control. For these reasons, RAT can still represent a useful resource in the context of massive screening among outpatients, providing that such assays would not be used in subjects with more than 4 days of symptoms and in subjects considered vulnerable. Moreover, we think that during epidemic waves, antigen tests may also prove to be useful at hospitals’ emergency rooms for patients’ cohorting, especially when rapid RT-PCR reagents are not available in sufficient numbers due to reagent shortage.

In order to compensate the lower sensitivity compared to RT-PCR, a paradigm change in RAT testing has been suggested (particularly in outpatients contexts), by increasing its frequency with the purpose of catching a greater number of patients during their high VL infection phase^20^. At the same time, RAT cannot substitute but only complement the RT-PCR testing, because of the gap in detection left among those patients with a VL, high enough to be contagious but not high enough to be detected by antigen tests.

As mentioned above, our results showed lower performances of RAT compared to data from a previous study performed in Lausanne^8^. First, this is due to the different patient population admitted at the emergency ward of our hospital as compared to outpatients (lower viral load among hospitalized subjects). Second, the study performed in the outpatients’ clinic was done at the time of explosive growth in number of cases, with most subjects exhibiting a recent infection. Third, patients arriving at the emergency ward of our university hospital were probably sicker, often for a longer time and possibly already developed an immune response, leading more frequently to lung edema, radiological infiltrate and hypoxemia. The presence of mucosal IgA targeting SARS-CoV-2 surface antigens might have played a role, thus competing with RAT for the same target. Interestingly, in our study none of the 41 patients with a positive RAT showed the presence of IgA, reinforcing the hypothesis that the presence of IgA might constitute a target competitor with the antibodies used in the antigen assay.

Finally, the implementation of RATs in patient triaging in the emergency department allowed us to save time and diagnostic reagents (which is particularly useful in periods of shortage). Nevertheless, in such population, due to the lower sensitivity of antigen tests, it is very important to perform systematically a confirmatory RT-PCR. This was done using different molecular platforms. Even though we adapted Ct results with regression equations, still confidence intervals of calculated Ct could not be taken into consideration to give a precise cut-off, thus leaving a margin of inter-variability, which might have partially affected our results. Finally, because nasopharyngeal swabs were transported in a VTM to be able to perform both RAT and RT-PCR analyses on the same sample, we hypothesized that the 2.5-3 ml dilution of the sample might have affected the sensitivity. To verify this hypothesis, we started a (still ongoing) prospective study comparing dry swabs (RAT immediately done at bedside) and wet swabs (RAT done in the laboratory, testing swabs put in the VTM). This ongoing clinical trial is showing so far exact same performances of the antigen test using both approaches, thus suggesting that a possible dilution effect is compensated by improved biological material release when the swab is immerged in the viral transport medium. Moreover, we observed also a very low sensitivity of 28% (even lower than the 33% sensitivity observed in the present study) with the same Standard Q antigen test among a cohort of subjects without COVID-19 symptoms, hospitalized in another hospital, despite their use of a dry swab approach (Caruana *et al*, submitted).

In conclusion, by keeping in mind the huge gap in technical sensitivity between RAT and RT-PCR (roughly corresponding to about 10.000 fold reduced analytical sensitivity), RAT might represent a valuable complementary tool, especially during outbreaks, when patient flow to the emergency department is particularly high and early orientation and effective cohorting is crucial.

## Data Availability

The data that support the findings of this study are not publicly available due to privacy restrictions.

## Contribution

Dr. Caruana conceived and design the study, performed the literature research, hands-on conduct of patients’ data collection and statistical analyses, drafted and revised the article. Dr Croxatto contributed to the design of the study, performed hands-on conduct of RAT validation and provided essential reagents and materials for the study. Dr. Carron contributed to the design of the study and provided essential reagents and materials for the study. Dr. Kampouri performed hands-on conduct of patients’ data collection. Dr. Kritikos contributed to patients’ data collection and statistical analyses. Dr. Opota, Dr. Foster, R. Brouillet, Dr. Senn provided essential reagents and materials for the study. Dr. Lienhard and Prof. Egli contributed to the conception and design of the study. Prof. Pantaleo performed hands-on conduct of the experiments and provided essential reagents and materials for the study. Prof. Greub conceived and designed the study, provided essential reagents and materials for the study and contributed to drafting the article. All authors critically revised the article and finally approved the version to be submitted.

## Acknowledgements

We would like to thank Gizha Shklqim for the precious help in the RAT laboratory set-up in the emergency ward. We also would like to thank all the staff of the Institute of Microbiology of the Lausanne University Hospital including all the biomedical technicians of the molecular diagnostic laboratory for routine RT-PCRs.

## Supplementary material

**Supplementary table 1.**
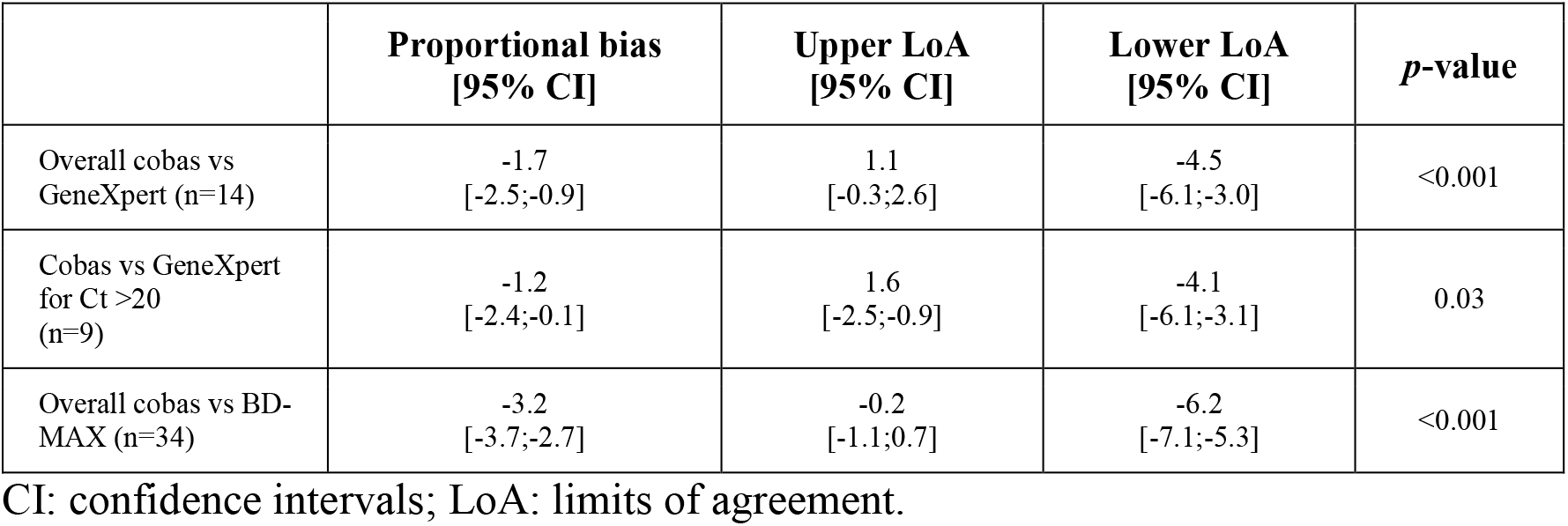
Bland-Altman analyses for inter-variability of Ct measure between cobas 6800 (Roche) versus GeneXpert (Cepheid) or BD-Max (Becton, Dickinson).

**Supplementary table 2.** RAT sensitivity rates after stratification for viral load.

| **RAT** | **Sensitivity among patients with VL>10<sup>5</sup>**<br>**(n=71)** | **Sensitivity among patients with VL>10<sup>6</sup>**<br>**(n=46)** | **Sensitivity among patients with VL>10<sup>7</sup>**<br>**(n=38)** |
| --- | --- | --- | --- |
| One Step | 74.6% | 100% | 100% |
| Standard Q | 66.2% | 97.8% | 100% |
| Panbio | 66.2% | 96.6% | 100% |
| BD Veritor | 64.8% | 95.6% | 100% |

**Supplementary figure 1.**
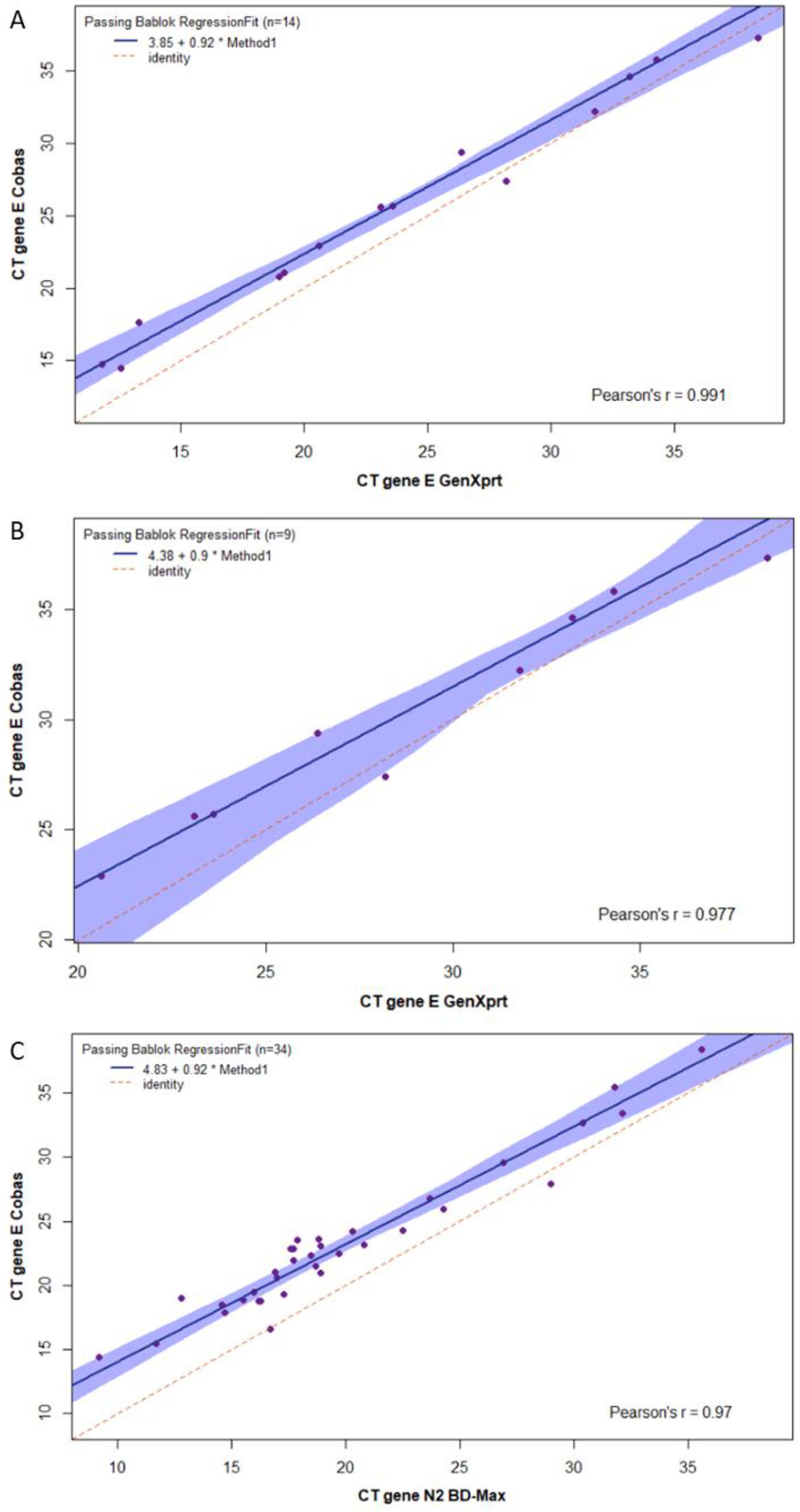
Passing-Bablock regressions for cobas 6800 Ct gene E against: A. GeneXpert gene E (for Ct >20); B. GeneXpert gene E (for Ct <20); C. BD-MAX gene N2.

**Supplementary figure 2.**
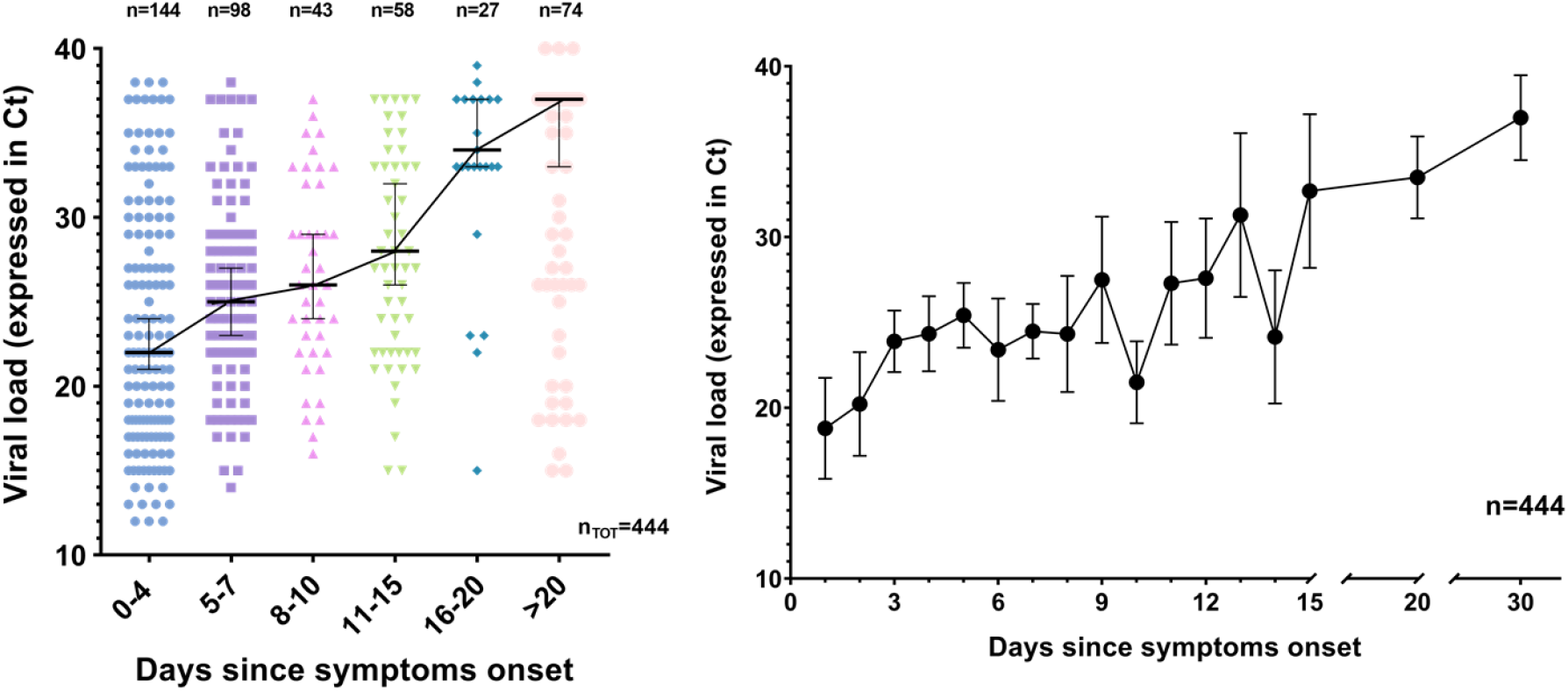
Changes of viral load over time (January-June 2020) according to symptoms delay.

